# A Cross-Sectional Evaluation of Health Resource Use in Patients with Functional Neurological Disorders Referred to a Tertiary Neuroscience Centre

**DOI:** 10.1101/2023.09.02.23294977

**Authors:** Brian O’Mahony, R. Nelson-Sice, Glenn Nielsen, Rachel Hunter, Sarah Cope, Niruj Agrawal, Mark Edwards, Mahinda Yogarajah

## Abstract

**Introduction:** Functional Neurological Disorder (FND) is a common cause of referral to neurology services. FND has been shown to lead to significant healthcare resource use, and is associated with significant disability, comorbidity, and distress. This leads to substantial direct, indirect, and intangible costs to the patient and society.

**Methods:** We recruited consecutive patients with FND referred to a tertiary FND specialist clinic. We assessed health and social care resource use, in the 6 months preceding their consultation through a modified version of the Client Service Receipt Inventory (CSRI) in the form of a postal questionnaire. The total cost was estimated by combining the number and frequency of health resource use with standard national unit costs. We also assessed indirect costs such as informal care and loss of income.

**Results:** We collected data on 118 subjects. Patients with comorbid anxiety or depression had higher costs in the preceding 6 months, as did patients who had a longer duration of FND symptoms. Indirect costs were larger than the already substantial direct costs, and a large proportion of FND patients were receiving government support.

**Conclusion:** This study highlights the high cost of FND to both patients and health systems. Adequate reform of the patient pathway and re-organization of services to make diagnoses, and initiate treatment more quickly, would likely reduce these costs.

## Introduction

Functional Neurological Disorder represent genuine involuntary neurological symptoms and signs which have characteristic clinical features and represent a problem of voluntary control and perception despite normal basic structure of the nervous system [1]. Manifestations of FND are varied, such as decreased or increased movement, loss of sensation, difficulties in speech, abnormal gait/posture, and seizure-like episodes (functional seizures) [FS]) [1]. FNDs can create a significant impact on their sufferer’s quality of life [2, 3], and patients often present with comorbid psychiatric conditions, with both depression and anxiety occurring in up to 40% of FND patients [4, 5].

FND of movement and sensation has a prevalence of roughly 50 per 100,000 population, and an incidence of 4–12 per 100,000 population per year. PNES contributes a further 1.5–4.9 per 100,000 population per year, with a prevalence of 2–33 per 100,000 population [6]. Patients with FND make up 9% of neurology admissions [7, 8], and 16% of neurology clinic referrals [9]. Delayed diagnoses of FND leads to worse outcomes for patients [4], as well as preventable costs, such as missed work, GP and specialist appointments, investigations etc. Diagnostic uncertainty in the midst of on-going symptoms can lead to intangible costs, such as decreased Quality of Life (QOL). These costs carry a burden to patients, clinicians, healthcare systems, and the economy.

The costs of FND (and other medical conditions) can be separated into direct and indirect costs. Direct costs represent resources utilised for health care (e.g. cost of investigations, time spent on assessment by a doctor etc), as well as out-of-pocket costs to the patient. Indirect costs represent productivity losses arising from morbidity-related sickness absence (e.g. loss of employment, cost of childcare while hospitalised etc.). Direct and indirect costs together constitute the economic burden of FND, which can be estimated by measuring the monetary valuation of health care utilisation and lost productivity in patient samples.

The literature concerning the economic cost of FND is sparse, and any conclusions which may be drawn from it are limited by the heterogeneity of the studies which focus on the topic. Studies vary in the costs included in their analysis, with many focussing only on hospital costs [10-14]. However, Stephen et al’s comprehensive study highlights that people with FND accrue similar costs to those with refractory epilepsy and demyelinating disorders. The cost of FND alone was estimated to be $1.2 billion annually in the United States of America in 2017 [13]. In Denmark, Jennum et. al showed a near tenfold increase of combined direct/indirect costs in FS patients compared to healthy controls [15]

Three studies [15-17] have reported on the cost of productivity loss due to FND. Each of these studies reported these costs as being higher than the direct medical costs resulting from the disorder. It has been found that FND patients are more likely not to be working for health reasons and more likely to be receiving disability-related state financial benefits than patients with other neurological patients [18]. No study has yet assessed whether symptom severity and/or duration impact the economic cost of FND.

In this study we set out to evaluate direct and indirect costs associated with FND through a retrospective questionnaire-based assessment of people referred to a tertiary FND specialist assessment clinic.

## Methods

### Participants and Setting

Participants were patients with scheduled new appointments at St George’s Hospital FND Clinic from 17^th^ October 2017 until 6^th^ February 2018. St George’s Hospital Neurology Department is the regional specialist tertiary neuroscience inpatient and outpatient centre for over 3 million people across South-West London, Surrey and Sussex.

Patients attending the clinic for follow up appointments and patients with primary diagnoses other than FND were excluded from the study. Diagnosis of FND was made by __________

### Data and Collection

Prior to attending their new appointment, patients were asked to complete a questionnaire. The questionnaire asked participants to retrospectively assess their NHS resource use including inpatient, outpatient, and community-based care. Patients were also asked to report the effect of FND on their own economic status, for example any change in employment and/or government benefits received. The study was registered and approved after review as a service evaluation with the clinical governance and audit office at St George’s Hospital.

Costs were measured in 2018 Pound Sterling (sign: £)

### Research Instruments

#### Client Service Receipt Inventory (CSRI)

A modified version of the Client Service Receipt Inventory (CSRI) was employed to quantify the health and social care resource use in the 6 months preceding patient consultation (see Appendix A). The CSRI has been used to quantify health and social care resource use in patients with chronic neurological disorders [19] and has displayed its reliability in obtaining an accurate inventory of data through which costs can be calculated. The CSRI was modified and adapted to be more specific to the cohort of FND patients based on previous CSRI-included studies [19] and informed input from specialist consultants in the FND clinic.

Healthcare resource data obtained by the modified CSRI included hospital outpatient appointments, treatments and medications, investigatory procedures, inpatient and residential care, and care provided by all primary and secondary healthcare professionals. Economic and social information included patient employment and informal care received by friends and relatives.

#### EuroQol-5D (EQ-5D)

The EQ-5D is a standardized instrument used for measuring generic health status. It is a self-reported scale, comprising of five dimensions; Mobility, Self-care, Usual activities, Pain/discomfort, and Anxiety/depression. Use of the EQ-5D aimed to investigate the relationship between symptomology and resource use in the cohort, more specifically that of symptom severity with frequency and type of resources used.

### Data Analysis

To provide an estimate of the cost of health and social care resource use, the type and frequency of resource use was combined with the national unit costs. National unit costs were extracted from “Unit costs of Health and Social Care 2015” [20] and supplementary sources [21, 22]. Medication costs were calculated using information in the British National Formulary [23].

When accounting for the contact time of the participants with the unit cost for specific health care professional, the patients’ account of contact time was deemed more reliable than the published “average consultation time”, based on the likelihood that the FND patient group deviate from the mean consultation time of all patients. The complexity of FND patients requires a multi-faceted consultation approach to address not only the physical symptoms, but also the psychological and social implications of FND. Therefore, use of “average consultation time” in this cohort would likely result in inaccurate patient costs [24].

The participants’ loss of employment income because of their FND were costed based on the employment income before and after symptom presentation based on data gathered in the CSRI ‘Section 2’. The value of income lost was estimated using national average salaries in line with the participant’s job sector and job title [25].

The informal care received by the participants was quantified using the replacement cost method [26], i.e. time spent by friends and relatives providing informal care and assistance was valued as equal to the cost of a paid professional that the friend and/or relative had hypothetically replaced. Therefore, the informal care received was valued at £18 per hour, equal to the Curtis 2015 data on a local authority care worker [20].

Statistical analysis was performed with JASP statistic software package. Data were expressed as means ± standard deviation (SD). Comparisons between groups were performed with analysis of non-parametric test. A value of P < 0.05 was considered statistically significant.

## Results

### Study demographics

Questionnaires were sent to 328 patients and were completed by 118 participants, a response rate of 36%. 83 questionnaires had every section completed. Most patients identified as White British (77%), followed by Black African and Black Caribbean (6.2% each).

### Direct health costs

Breakdown of costs by service are given in Tables 1 and 2. Despite being used by only 6.36% of patients, the cost associated with Intensive Care Unit admissions had the highest mean cost per patient at £629.15. This was followed by neurology ward admissions (also utilised by only 6.36% of patients) which carried a mean cost of £245.38. General Practitioner consultations, whether in person (81% of patients) or by phone (46.36% of patients), were used by most patients, and carried a mean cost per patient of £245.

**Table 1:**
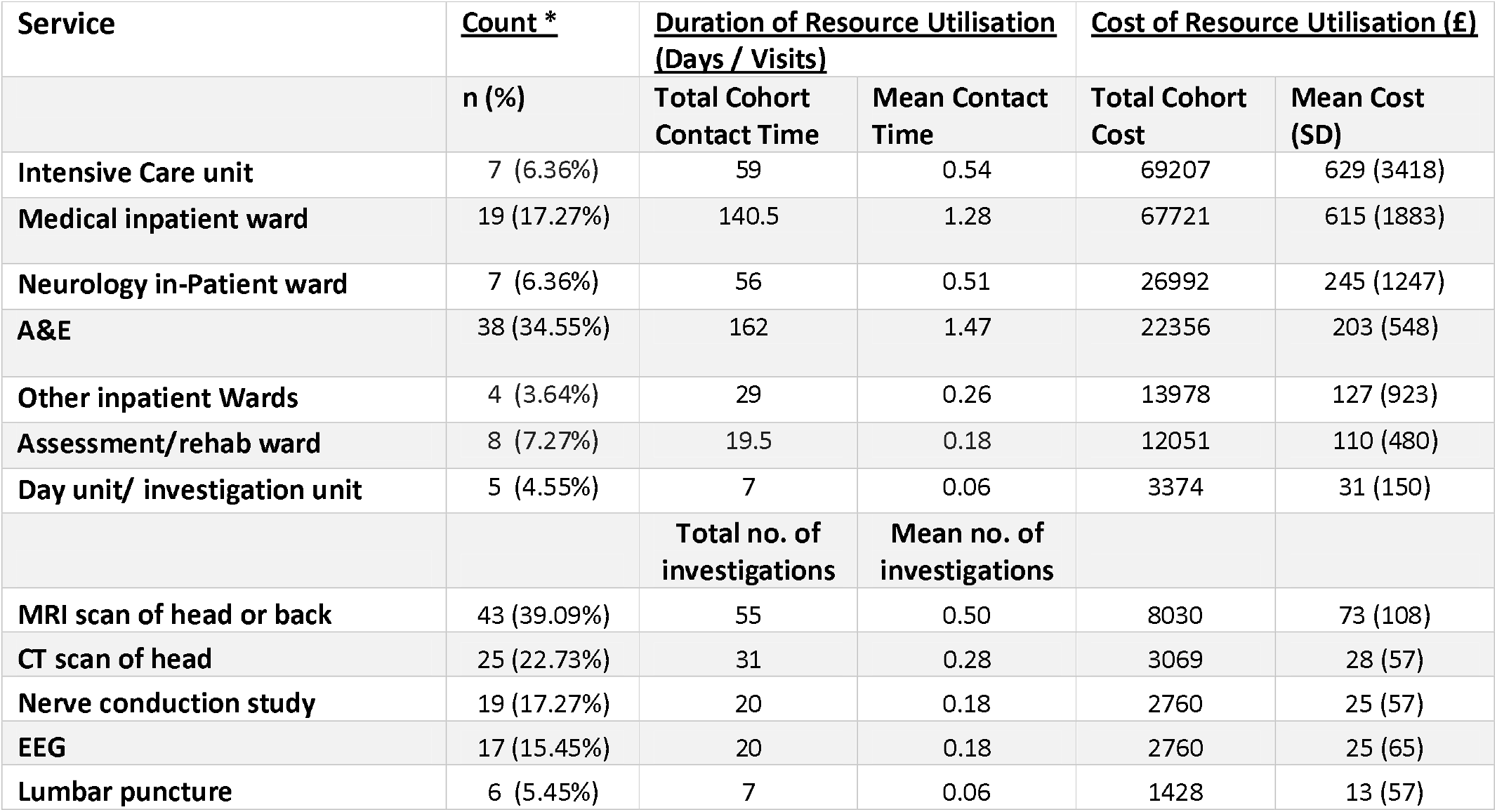
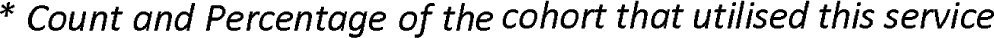
Inpatient Service utilisation and cost in the 6 months prior to new appointments (n=110)

Count and Percentage of the cohort that utilised this service
| Service | Count * | Duration of Resource Utilisation<br>(Days / Visits) |  | Cost of Resource Utilisation (£) |  |
| --- | --- | --- | --- | --- | --- |
|  | n (%) | Total Cohort<br>Contact Time | Mean Contact<br>Time | Total Cohort<br>Cost | Mean Cost<br>(SD) |
| Intensive Care unit | 7 (6.36%) | 59 | 0.54 | 69207 | 629 (3418) |
| Medical inpatient ward | 19 (17.27%) | 140.5 | 1.28 | 67721 | 615 (1883) |
| Neurology in-Patient ward | 7 (6.36%) | 56 | 0.51 | 26992 | 245 (1247) |
| A&E | 38 (34.55%) | 162 | 1.47 | 22356 | 203 (548) |
| Other inpatient Wards | 4 (3.64%) | 29 | 0.26 | 13978 | 127 (923) |
| Assessment/rehab ward | 8 (7.27%) | 19.5 | 0.18 | 12051 | 110 (480) |
| Day unit/ investigation unit | 5 (4.55%) | 7 | 0.06 | 3374 | 31 (150) |
|  |  | Total no. of<br>investigations | Mean no. of<br>investigations |  |  |
| MRI scan of head or back | 43 (39.09%) | 55 | 0.50 | 8030 | 73 (108) |
| CT scan of head | 25 (22.73%) | 31 | 0.28 | 3069 | 28 (57) |
| Nerve conduction study | 19 (17.27%) | 20 | 0.18 | 2760 | 25 (57) |
| EEG | 17 (15.45%) | 20 | 0.18 | 2760 | 25 (65) |
| Lumbar puncture | 6 (5.45%) | 7 | 0.06 | 1428 | 13 (57) |

**Table 2:**
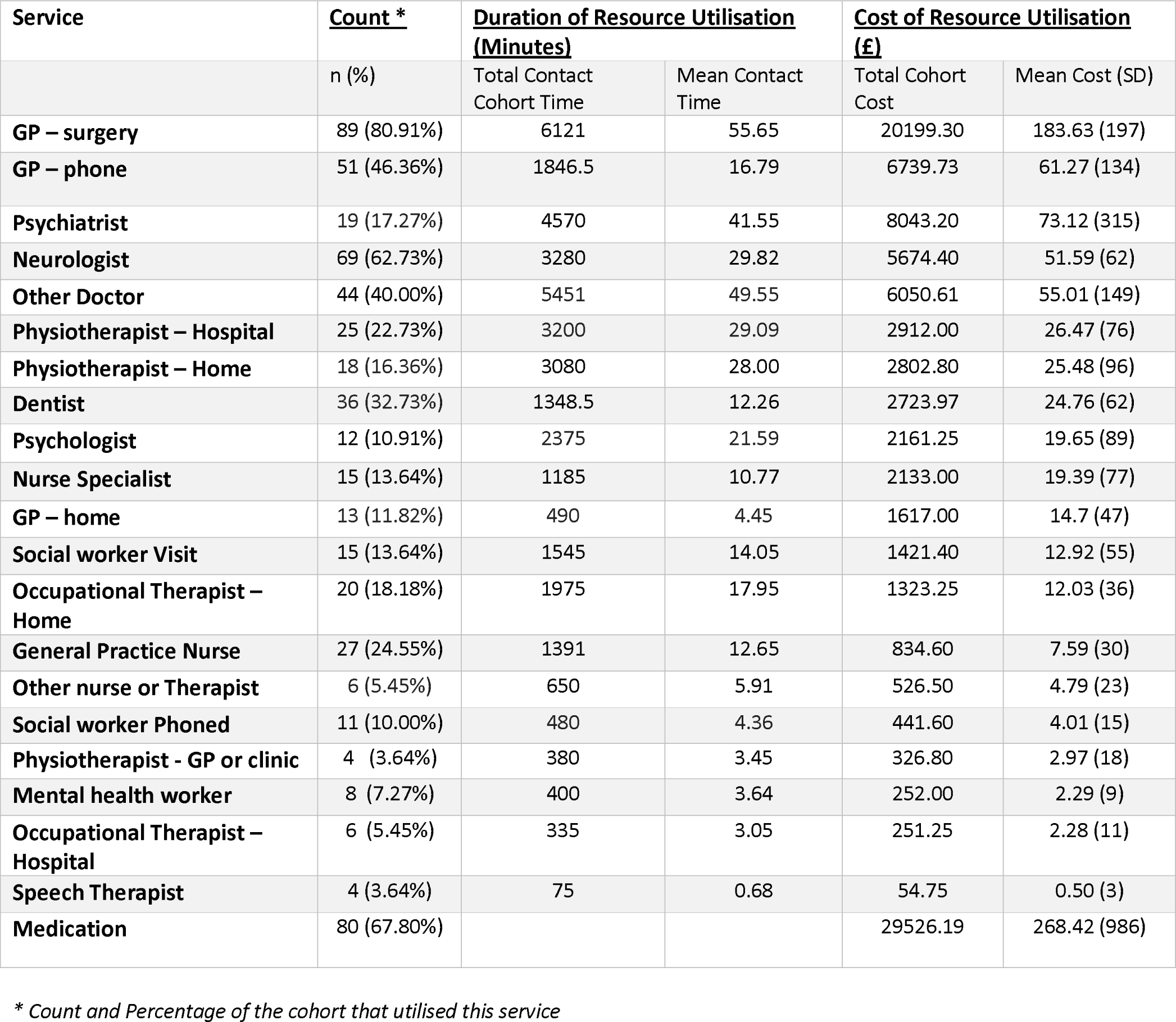
Outpatient Service utilisation and cost in the 6 months prior to new appointments (n=110)

Of note patients who were depressed (defined as PHQ9 score of >10) incurred greater mean costs to the NHS than those patients who were defined as not depressed (£4380 vs £1503, t(83) = -3.25, p<0.001). The same phenomenon was true for patients defined as anxious (GAD7 score of >10) vs those defined as not anxious (£4,017 vs £1,980, t(82) = -2.1, p<0.001).

There was no significant relationship between duration of disorder and EQ-5D score (p=0.36, Spearman’s r=0.13). After removal of outliers, defined as >3 standard deviations from the mean, there was a significant relationship between duration of symptoms and total cost to NHS in the prior 6 months (p=0.04, Spearman’s r =.226).

Many participants incurred substantial out-of-pocket expenses in the form of adaptations made to residences for the purpose of disabled access. 26.76% of participants made adaptations for disabled access primarily to lavatory facilities and kitchens. The mean out-of-pocket expense of these participants who made modifications was £3499.47 (±£5299.60) while the mean across the full cohort was £570.85 (±£2446.71).

Home based services are summarised in Table 3. A small minority of patients utilised these services, which perhaps highlights the skewed distribution of the health-resource use of FND patients.

**Table 3:**
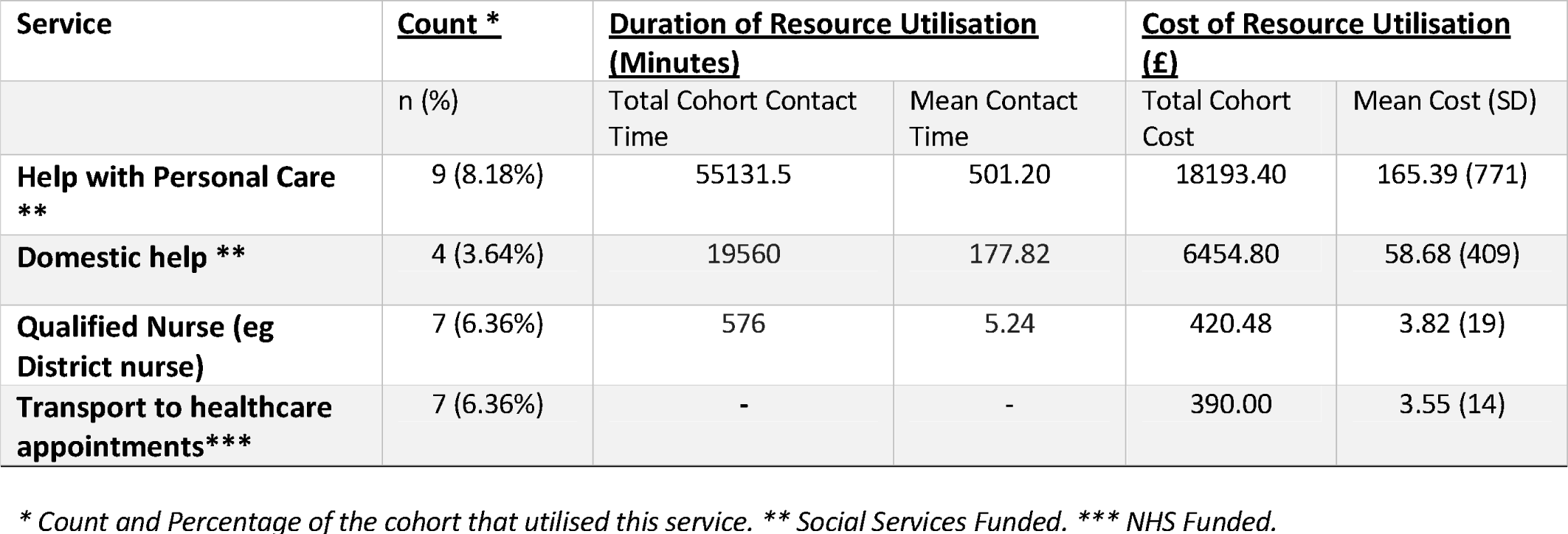
Utilisation of Home-Based Services and cost in the 6 months prior to new appointments (n=110)

Total costs to the NHS by patient are displayed in figure 1. These costs include all costs listed in Tables 1 and 2, as well transport to NHS appointments and visits by the district nurse. The mean cost per patient was £3,229 (95% Confidence Interval of £2220 - £4240), with a median value of resource use of £1,152.27. 12.68% of respondents reported costs of over £5000, predominantly due to in-patient admission.

**Figure 1:**
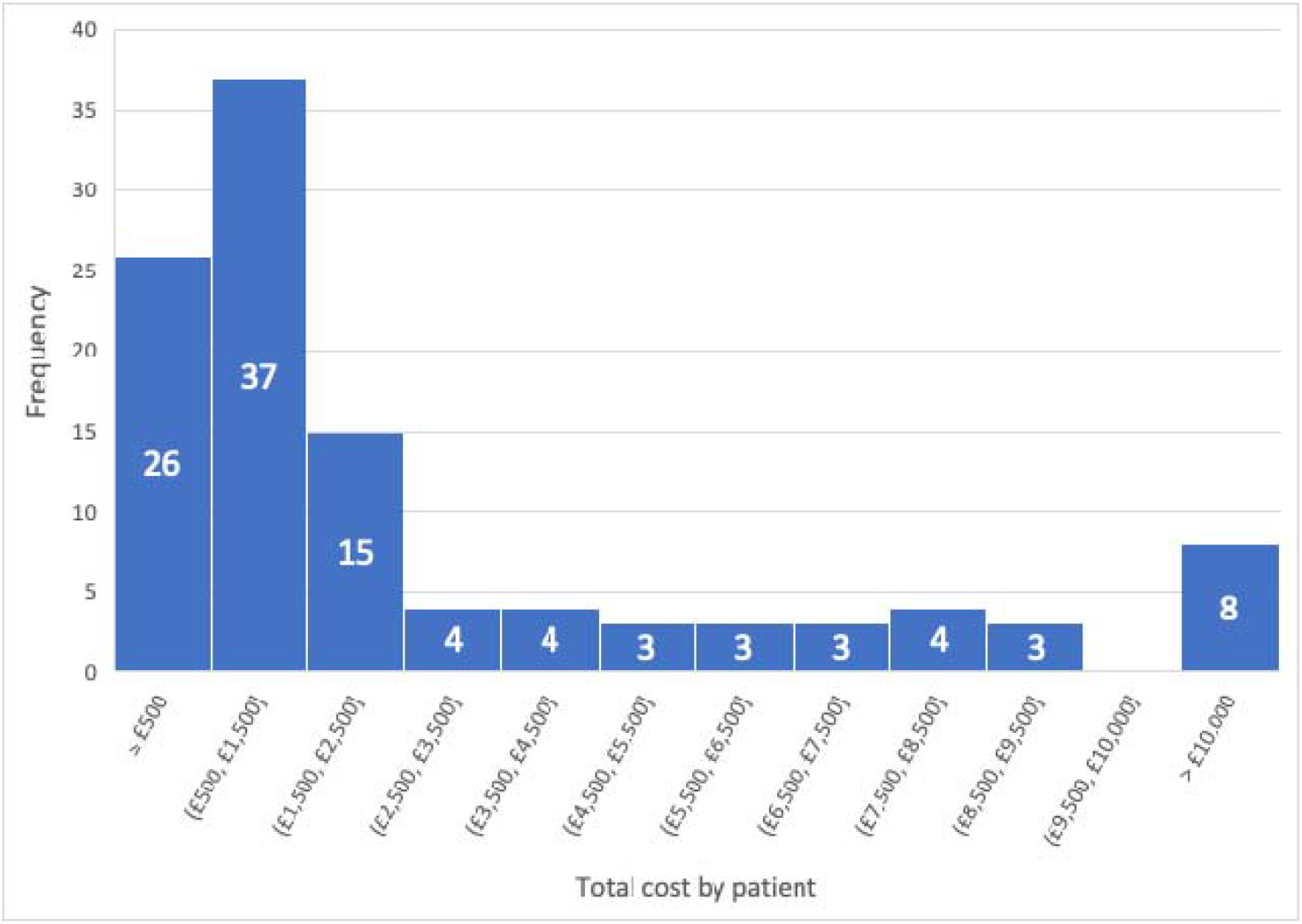
Costs to NHS by patient in the 6 months prior to new appointments

### Indirect costs

There was a substantial cost of lost income in the cohort, which was calculated as estimated annual income prior to the onset of their FND, less annual income after onset. (Table 4), estimated at a total of £758,355 among 115 patients. This represents a mean of £6594.4 (SD=8503) amongst all patients. Excluding participants who were unemployed prior to symptom onset mean loss of income was £10 821 91 (±8306) Figure 2 shows the amount of Only 16.5% of study participants studied were able to maintain full-time employment, with another 10.4% employed on a part-time basis. With their lack of income from employment, many patients became reliant on government benefits to supplement or replace income. Of the cohort, 71.8% received welfare benefits over the preceding 6 months, the mean amount of received being £299.50 (±180.76) per week Loss-of-productivity affected not only patients, but also their carers, friends, and family as shown in table 5. Patients estimated receiving a mean of almost 20 hours per week (median 13.75 hours per week) of informal care.

**Table 4:**
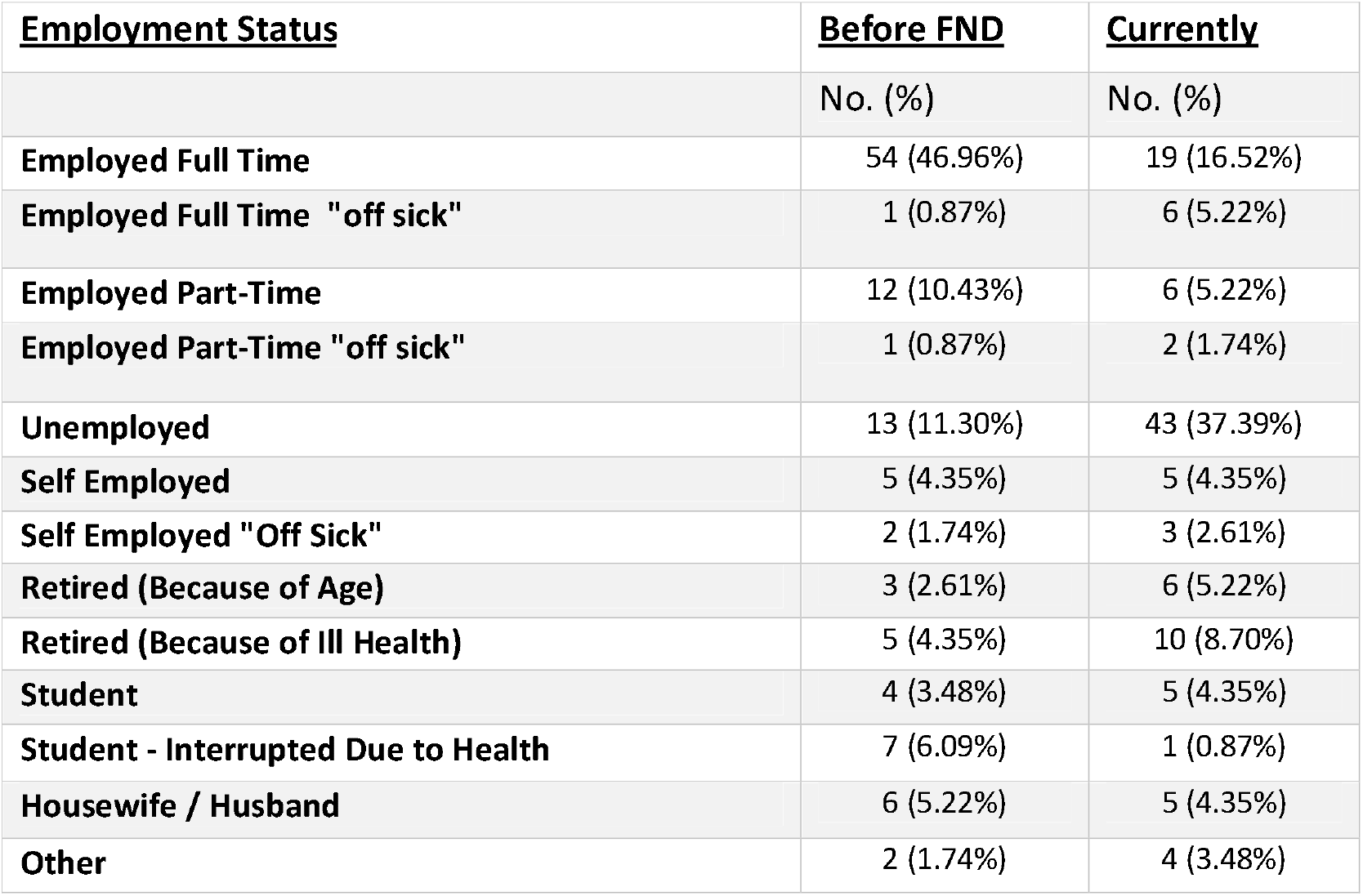
Change in Employment Status due to their FND (n = 115)

| **<u>Employment Status</u>** | **<u>Before FND</u>** | **<u>Currently</u>** |
| --- | --- | --- |
|  | No. (%) | No. (%) |
| **Employed Full Time** | 54 (46.96%) | 19 (16.52%) |
| **Employed Full Time "off sick"** | 1 (0.87%) | 6 (5.22%) |
| **Employed Part-Time** | 12 (10.43%) | 6 (5.22%) |
| **Employed Part-Time "off sick"** | 1 (0.87%) | 2 (1.74%) |
| **Unemployed** | 13 (11.30%) | 43 (37.39%) |
| **Self Employed** | 5 (4.35%) | 5 (4.35%) |
| **Self Employed "Off Sick"** | 2 (1.74%) | 3 (2.61%) |
| **Retired (Because of Age)** | 3 (2.61%) | 6 (5.22%) |
| **Retired (Because of Ill Health)** | 5 (4.35%) | 10 (8.70%) |
| **Student** | 4 (3.48%) | 5 (4.35%) |
| **Student - Interrupted Due to Health** | 7 (6.09%) | 1 (0.87%) |
| **Housewife / Husband** | 6 (5.22%) | 5 (4.35%) |
| **Other** | 2 (1.74%) | 4 (3.48%) |

**Figure 2:**
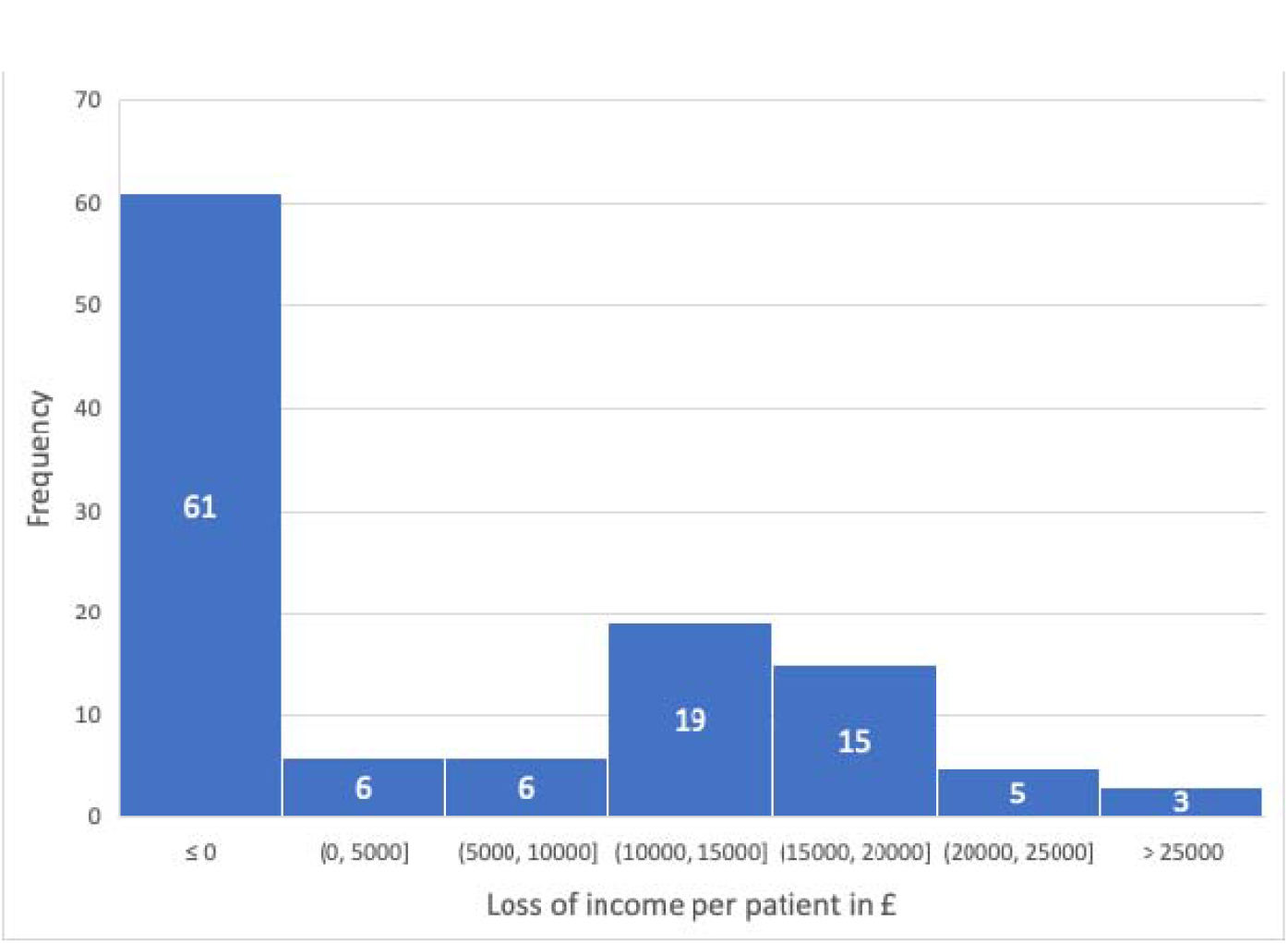
Annual income loss per patient due to their FND

**Table 5:** Weekly hours and Cost of informal Care (n = 100)

| **<u>Informal Care Method</u>** | **<u>Mean hours spent per week (SD)</u>** | **<u>Mean estimated Value per week (SD)</u>** |
| --- | --- | --- |
| **Personal Care** | 3.16 (5.64) | 56.88 (102) |
| **Housework** | 4.11625 (6.12) | 74.0925 (110) |
| **Transport** | 2.9175 (3.58) | 52.515 (65) |
| **Preparing meals** | 3.46 (4.96) | 62.28 (89) |
| **Gardening** | 0.7125 (1.25) | 12.825 (23) |
| **Shopping** | 1.72 (2.28) | 30.96 (41) |
| **Looking after pets** | 1.63 (3.87) | 29.34 (70) |
| **Home improvements** | 0.81 (1.68) | 14.58 (30) |
| **Other** | 1.25 (8.16) | 22.5 (147) |
| **Total Informal Care** | 19.78 (21.89) | 355.97 (394) |

### Total health costs

Table 6 shows total health costs of the cohort. Total costs were also positively skewed, with a skewness value of 0.78, and kurtosis value of -0.719.

**Table 6:** Summary of 6-month costs by service type (n = 118)

|  | **Mean\* (SD)** | **Median\*** | **Range** |
| --- | --- | --- | --- |
| **Inpatient Service Use** | 1960.72 (4560.89) | 0 | 0-33396 |
| **Outpatient** | 604.45 (596.56) | 431.9 | 0-3015 |
| **Medication** | 268.42 (986.30) | 50.1 | 0-10089.64 |
| **Home based services** | 231.44 (876.017) | 0 | 0-4752 |
| **Diagnostic Investigations** | 164.06 (211.56) | 138 | 0-1196 |
| **Employment Lost** | 6594.40 (8503.74) | 0 | 0-30048 |
| **Informal Care** | 9255.29 (10242.27) | 6435 | 0-41184 |
| **Adaptations** | 570.85 (2446.71) | 0 | 0-18200 |
| **Total Service Cost** | 3229.09 (5395.93) | 1117.54 | 0-34915.64 |

## Discussion

This study highlights high rates of health resource utilisation in patients with FND. These patients were found to have a mean utilisation of health resources valued at £3,229 over the 6-month period (£6,458 p/a) prior to their initial appointment at the tertiary neuropsychiatry service. Extrapolation of this mean value using an estimated incidence of 4-12 per 100,000 per year [27-29] gives a total cost of NHS resource use of between £13.5 million and £40.4 million per year. This estimate is nearly eight times that of the national health expenditure per person, almost twice that of estimates of the cost of Chronic Obstructive Airways Disease (£3,488 p/a) and is almost four times that of depression (£1,873 p/a) and diabetes (1,870 p/a) [30]. Such comparisons are, however, limited by heterogenous methods of cost-estimation. Jennum et al [15] compared the cost of PNES patients to age- and location-matched controls and found direct healthcare costs to be 4.8 times greater in the PNES group. Both findings demonstrate the high direct healthcare costs of people with FND.

The distribution of these direct costs was positively skewed, resulting from a small number of patients requiring the use of costly interventions, including admissions to hospital and intensive care units. The most frequently used services were outpatient services, particularly General Practitioners. However, as shown in Tables 1 and 2, the most-costly resource utilisations in the cohort were, in decreasing order, admission to an intensive care unit, admission to a medical in-patient ward, general practitioner appointments, and emergency department visits.

As in other cost of illness analyses, it is difficult to isolate the “pure” cost of FND, i.e. that cost which does not stem from any comorbid conditions. Any cost estimates reported in this study represent the yearly direct and indirect costs of patients with FND. Attempting to assess such a pure cost may be an exercise in futility, given the nature of the interaction of FND with its psychiatric comorbidities. Whether FND symptom severity and outcome is positively or negatively affected by a mood or anxiety disorder is unclear [5, 31-33]. Our findings suggest that, in any case, symptom severity is not correlated with higher health costs. Our findings of increased costs of FND patients who also suffer from depression and anxiety may indicate only an added cost of these two disorders, which has been described in the literature [34, 35], or it may point to a synergistic relationship. The investigation of this question is beyond the scope of a self-reported, retrospective review, but may offer an avenue for future research.

As is the case in previous studies which investigated indirect costs of FND [15-17], our findings show that the indirect costs of the disorder dwarf the already considerable direct costs. Total indirect costs per patients were a mean of £15,850. Such indirect costs are borne by both patients and their family/friends, as well as by the taxpayers in supporting those no longer able to gain money from employment. Such high indirect costs are compounded by FND patients having worse outcome. when in receipt of government welfare [36].

Comparing our findings to the literature of economic costs of FND is challenging given the geographical, clinical, and methodological heterogeneity of the studies in this area. Looking specifically at studies investigating adults with FND in countries with a similar, public healthcare system; In patients with PNES, Goldstein et al [16] in the United Kingdom found similar health care utilisation costs (£3,943), but substantially lower productivity loss (£2,953) in a cohort of n=367, in the six months prior to treatment. Magee et al [17] in Ireland assessed the cost of PNES to tax-payers, and reported direct costs of €2,714.5 per 6 months, with combined social welfare payments and loss of tax revenue costs calculated at €7,783 per 6 months per person. Deleuran et al in Denmark found direct healthcare costs €2,904 over 6 months in PNES. Finally, Tinazzi et al in Italy reported an average direct hospital cost of €1,652 per 6 months in patients referred to a specialist Functional Movement Disorder clinic.

### Implications for clinical practice

An important finding from the literature relating to treatment interventions in FND is a decline in healthcare resource utilisation [37-39] and economic cost [14, 16, 40-43] following an intervention, whether this is psychotherapy, structured delivery of the diagnosis or specialist physiotherapy. Our findings suggest that patients with a longer duration of FND continue to have higher costs in the months prior to diagnosis than those with shorter duration. Additionally, our findings suggest that it is incorrect to assume that the correlation between NHS resource use and duration of disorder is a result of reduced quality of life. This highlights the potential cost saving of early intervention to minimise monetary and quality of life costs to both the patient and society. An important first step would be to increase patient access to specialist services and/or to improve general knowledge of the condition. Referral to an FND specialist may reduce the latency to diagnosis and avoid unnecessary consultations and tests [28]. This study investigated only costs in the six months prior to the patients’ attendance at the FND clinic, and costs subsequent to this should be studied to investigate any change. Given that availability of adequate treatment of FND is limited, one might expect chronicity of the disorder and thus ongoing, long term costs of A&E visits, investigations, and admissions, as well as indirect costs.

### Limitations

We acknowledge some study limitations. Firstly, use of the patient-reported CSRI is liable to recall bias. While this limitation should be considered, the results of a 2005 study suggest that retrospective self-report data can be equally reliable as medical database data [44]. Also of note, data gathered on participants’ medication use through self-reporting had a surprisingly low completion rate. Rather than this signifying that fewer patients than expected were taking medication, it could be that participants did not complete this section due to lack of knowledge of the name of their medications. Therefore, one can argue the data on medication should be treated as a minimum possible value.

A second limitation is the low return rate of the questionnaire, resulting in a relatively small sample size. Furthermore, the large number of questionnaire non-responders (63.99% of the intended cohort) could signify a selection bias in the study, limiting the external validity of these findings.

Thirdly, the patients included in our study are those referred to a tertiary specialist service, and as such likely represent a severely affected cohort. Such referral bias would also limit the external validity of the study’s results.

Lastly, our study would ideally have utilised a comparator group, so that the costs associated with FND could have been contextualised. Failing use of a comparison with healthy controls or another neurological disorder, one possibility might have been to compare the costs of patients before and after their diagnosis of FND, as there has yet to be a study comparing indirect costs before and after patients receive a diagnosis of FND.

## Conclusions

This study highlights the high cost of FND to both patients and the NHS. Patients with a longer duration of suffering from FND, were shown to have higher cost in the preceding 6 months. Our findings are consistent with similar studies’ reporting of the high direct, and higher still indirect cost of the disorder. Adequate reform of the patient pathway and re-organization of NHS services to make diagnoses and initiate treatment more quickly would likely reduce these costs.

## Data Availability

All data produced in the present study are available upon reasonable request to the authors

## References

1. Espay, A.J., et al., Current concepts in diagnosis and treatment of functional neurological disorders. JAMA neurology, 2018. 75(9): p. 1132–1141.

2. Věchetová, G., et al., The impact of non-motor symptoms on the health-related quality of life in patients with functional movement disorders. Journal of psychosomatic research, 2018. 115: p. 32–37.

3. Gelauff, J.M., et al., Fatigue, not self-rated motor symptom severity, affects quality of life in functional motor disorders. Journal of Neurology, 2018. 265(8): p. 1803–1809.

4. Gelauff, J., et al., The prognosis of functional (psychogenic) motor symptoms: a systematic review. Journal of Neurology, Neurosurgery & Psychiatry, 2014. 85(2): p. 220–226.

5. Feinstein, A., et al., Psychiatric outcome in patients with a psychogenic movement disorder: a prospective study. Cognitive and Behavioral Neurology, 2001. 14(3): p. 169–176.

6. Carson, A. and A. Lehn, Chapter 5 - Epidemiology, in Handbook of Clinical Neurology, M. Hallett, J. Stone, and Carson, Editors. 2016, Elsevier. p. 47–60.

7. Lempert, T., et al., Psychogenic disorders in neurology: frequency and clinical spectrum. Acta Neurologica Scandinavica, 1990. 82(5): p. 335–340.

8. Beharry, J., et al., Functional neurological disorders presenting as emergencies to secondary care. European Journal of Neurology, 2021. 28(5): p. 1441–1445.

9. Stone, J., et al., Who is referred to neurology clinics?—The diagnoses made in 3781 new patients. Clinical Neurology and Neurosurgery, 2010. 112(9): p. 747–751.

10. Seneviratne, U., et al., Medical health care utilization cost of patients presenting with psychogenic nonepileptic seizures. Epilepsia, 2019. 60(2): p. 349–357.

11. Luthy, S.K., et al., Characteristics of children hospitalized for psychogenic nonepileptic seizures due to conversion disorder versus epilepsy. Hospital Pediatrics, 2018. 8(6): p. 321–329.

12. Goyal, N., et al., Cost burden of stroke mimics and transient ischemic attack after intravenous tissue plasminogen activator treatment. Journal of Stroke and Cerebrovascular Diseases, 2015. 24(4): p. 828–833.

13. Stephen, C.D., et al., Assessment of emergency department and inpatient use and costs in adult and pediatric functional neurological disorders. JAMA neurology, 2021. 78(1): p. 88–101.

14. Russell, L.A., et al., A pilot study of reduction in healthcare costs following the application of intensive short-term dynamic psychotherapy for psychogenic nonepileptic seizures. Epilepsy & Behavior, 2016. 63: p. 17–19.

15. Jennum, P., R. Ibsen, and J. Kjellberg, Welfare consequences for people diagnosed with nonepileptic seizures: A matched nationwide study in Denmark. Epilepsy & Behavior, 2019. 98: p. 59–65.

16. Goldstein, L.H., et al., Cognitive-behavioural therapy compared with standardised medical care for adults with dissociative non-epileptic seizures: the CODES RCT. Health Technology Assessment (Winchester, England), 2021. 25(43): p. 1.

17. Magee, J., et al., The economic cost of nonepileptic attack disorder in Ireland. Epilepsy & Behavior, 2014. 33: p. 45–48.

18. Carson, A., et al., Disability, distress and unemployment in neurology outpatients with symptoms ‘unexplained by organic disease’. Journal of Neurology, Neurosurgery &amp; Psychiatry, 2011. 82(7): p. 810–813.

19. Jackson, D., et al., Service use and costs for people with long-term neurological conditions in the first year following discharge from in-patient neuro-rehabilitation: a longitudinal cohort study. PLoS One, 2014. 9(11): p. e113056.

20. Curtis, L.A. and A. Burns, Unit costs of health and social care 2015. 2015: Personal Social Services Research Unit.

21. Center, N.D.a.T. Cost Comparison Charts. 2017 October 2017]; Available from: http://gmmmg.nhs.uk/docs/cost_comparison_charts.pdf.

22. Care, U.D.o.H.a.S., National Schedule of reference costs: 2015-2016. 2016.

23. Committee, J.F., British National formulary 72 (September 2016-March 2017). London: British National Formulary, 2017.

24. Zantinge, E.M., et al., The workload of GPs: consultations of patients with psychological and somatic problems compared. British Journal of General Practice, 2005. 55(517): p. 609–614.

25. Smith, R., Employee earnings in the UK: 2018. Office for National Statistics, 2018.

26. Koopmanschap, M.A., et al., An overview of methods and applications to value informal care in economic evaluations of healthcare. Pharmacoeconomics, 2008. 26(4): p. 269–280.

27. Binzer, M., P.M. Andersen, and G. Kullgren, Clinical characteristics of patients with motor disability due to conversion disorder: a prospective control group study. Journal of Neurology, Neurosurgery & Psychiatry, 1997. 63(1): p. 83–88.

28. Stone, J., et al., Who is referred to neurology clinics?—the diagnoses made in 3781 new patients. Clinical neurology and neurosurgery, 2010. 112(9): p. 747–751.

29. Szaflarski, J.P., et al., Four-year incidence of psychogenic nonepileptic seizures in adults in Hamilton County, OH. Neurology, 2000. 55(10): p. 1561–1563.

30. Public Health England The health and social care costs of a selection of health conditions and multi-morbidities, P. Health, Editor. 2020.

31. Jalilianhasanpour, R., et al., Secure attachment and depression predict 6-month outcome in motor functional neurological disorders: a prospective pilot study. Psychosomatics, 2019. 60(4): p. 365–375.

32. Thomas, M., K.D. Vuong, and J. Jankovic, Long-term prognosis of patients with psychogenic movement disorders. Parkinsonism & related disorders, 2006. 12(6): p. 382–387.

33. Forejtová, Z., et al., The complex syndrome of functional neurological disorder. Psychological Medicine, 2022: p. 1–11.

34. Luppa, M., et al., Cost-of-illness studies of depression: A systematic review. Journal of Affective Disorders, 2007. 98(1): p. 29–43.

35. Revicki, D.A., et al., Humanistic and economic burden of generalized anxiety disorder in North America and Europe. Journal of affective disorders, 2012. 140(2): p. 103–112.

36. Gelauff, J. and J. Stone, Prognosis of functional neurologic disorders. Handbook of clinical neurology, 2016. 139: p. 523–541.

37. Nunez-Wallace, K.R., et al., Health resource utilization among US veterans with psychogenic nonepileptic seizures: A comparison before and after video-EEG monitoring. Epilepsy Research, 2015. 114: p. 114–121.

38. Razvi, S., S. Mulhern, and R. Duncan, Newly diagnosed psychogenic nonepileptic seizures: health care demand prior to and following diagnosis at a first seizure clinic. Epilepsy & Behavior, 2012. 23(1): p. 7–9.

39. Mayor, R., et al., Long-term outcome of brief augmented psychodynamic interpersonal therapy for psychogenic nonepileptic seizures: seizure control and health care utilization. Epilepsia, 2010. 51(7): p. 1169–1176.

40. Deleuran, M., et al., Psychogenic nonepileptic seizures treated with psychotherapy: long-term outcome on seizures and healthcare utilization. Epilepsy & Behavior, 2019. 98: p. 195–200.

41. Martin, R.C., et al., Improved health care resource utilization following video-EEG-confirmed diagnosis of nonepileptic psychogenic seizures. Seizure, 1998. 7(5): p. 385–390.

42. Ahmedani, B.K., et al., Diagnosis, costs, and utilization for psychogenic non-epileptic seizures in a US health care setting. Psychosomatics, 2013. 54(1): p. 28–34.

43. Chemmanam, T., et al., A prospective study on the cost-effective utilization of long-term inpatient video-EEG monitoring in a developing country. Journal of Clinical Neurophysiology, 2009. 26(2): p. 123–128.

44. Patel, A., et al., A comparison of two methods of collecting economic data in primary care. Family practice, 2005. 22(3): p. 323–327.

